# Physician- versus Large Language Model-Generated Clinical Summaries in the Emergency Department

**DOI:** 10.1101/2025.08.13.25333609

**Authors:** Niloufar Golchini, Nikita Mehandru, Ahmed Alaa, Melanie Molina

**Affiliations:** Department of Computational Precision Health, University of California, Berkeley and San Francisco, CA, USA; Bakar Computational Health Sciences Institute, University of California, San Francisco, CA, USA; Department of Emergency Medicine, University of California, San Francisco, California, USA; Division of Clinical Informatics and Digital Transformation, University of California, San Francisco, CA, USA

## Abstract

**Background:** As part of routine practice and documentation, emergency department (ED) clinicians routinely construct “one-liner” summaries—brief, information-rich statements distilling a patient’s history and presentation to support rapid decision-making. Producing these summaries is cognitively demanding and contributes to documentation burden. Large language models (LLMs) may assist by synthesizing longitudinal electronic health record (EHR) data.

**Methods:** We conducted a blinded, within-subject study of 99 ED encounters from March 2022–March 2024 at the University of California, San Francisco. We used an LLM to generate one-liner summaries using a k-nearest-neighbor few-shot prompting approach and clinical notes spanning multiple prior encounters. Twenty-one emergency physicians evaluated paired LLM- and physician-authored summaries in randomized order, rating accuracy, completeness, and clinical utility on 5-point Likert scales and indicating their overall preference with optional free-text explanation. Ratings were analyzed using linear mixed-effects models with summary type as a fixed effect and reviewer as a random intercept. Secondary analyses examined the LLM’s note-selection behavior and how inclusion of specific note types affected summary quality. We used rapid content analysis to review free-text explanations, identifying recurrent themes among reviewer preferences.

**Results:** Across all dimensions, LLM-generated summaries received higher ratings than physician-authored summaries. Mean (SE) estimated marginal means for accuracy were 4.18 (0.09) vs 3.40 (0.11) (β = 0.78; 95% CI 0.50–1.07; p < .001), for completeness 3.69 (0.10) vs 3.25 (0.12) (β = 0.44; 95% CI 0.14–0.74; p = .005), and for clinical utility 3.88 (0.10) vs 3.21 (0.12) (β = 0.67; 95% CI 0.35–0.99; p < .001). LLM-generated summaries were preferred in 50.5% of encounters, physician summaries in 38.4%, and 11.1% were rated equivalent. Qualitative analysis indicated that LLM summaries were often more inclusive and neutrally phrased, whereas physician summaries exhibited greater contextual nuance but occasionally omitted key details.

**Conclusions:** In this blinded evaluation of ED encounters, LLM-generated one-liner summaries outperformed physician-authored summaries on accuracy, completeness, and clinical utility. Patterns in note utilization suggest that models selectively integrate high-yield clinical sources, which may have important implications for the cost and efficiency in future healthcare deployment. These findings represent an important first step toward leveraging LLMs to aid rapid synthesis of complex EHR data in high-stakes environments.

## INTRODUCTION

Emergency physicians make rapid, high-stakes decisions amid persistent time pressures, resource constraints, frequent interruptions, and emergency department crowding.^1,2^ Upon first meeting a patient, they must quickly integrate clinical data from multiple sources within a patient’s electronic heath record (EHR) into a coherent mental model that supports diagnostic reasoning and immediate management. A key early step in this process is constructing a concise “one-liner”—an ED-specific clinical summary that distills the most important aspects of a patient’s demographics, medical history, and chief complaint into a single interpretive statement that typically opens the “History of Present Illness” section of the clinical note. Unlike clinical settings where physicians know their patients longitudinally, ED physicians encounter patients without prior familiarity, making the rapid synthesis and summarization of relevant clinical information especially critical for situational awareness. In addition to facilitating formal handoff and documentation, the ED one-liner functions as a cognitive scaffold: it concisely summarizes relevant clinical context and frames the patient’s presenting complaint.^3, 4^ Because the one-liner often represents the ED physician’s first cognitive synthesis of the patient’s clinical history and presentation, its quality and accuracy can shape initial diagnostic reasoning and influence subsequent decisions.^5^

Crafting a high-quality one-liner is cognitively demanding. On average, ED physicians are interrupted 12 to 13 times per hour, making navigating complex longitudinal records authored by multiple providers across varied care settings a challenging task.^6^ These records often contain diverse note types (e.g. discharge summaries, consult notes, imaging reports) from different institutions, requiring ED physicians to search for and filter through specific pieces of information.^7^ Such cognitive labor competes directly with bedside patient care and contributes to fatigue and burnout.^8^ Yet the cost of omission or misrepresentation in an ED one-liner can be clinically consequential, particularly when decisions must be made under time pressure and with limited information.^9^

Large language models (LLMs) have recently shown strong performance in clinical reasoning and summarization tasks across a range of benchmarks and use cases.^10–12^ Clinical applications have focused on generating long-form summaries, such as hospital discharge summaries, using notes from single encounters as inputs.^13–16^ However, no study to date has systematically evaluated the ability of LLMs to generate concise “one-liner” summaries, which would require integrating multiple EHR information types (e.g., clinical notes, imaging reports) and clinical encounters across time. This gap is particularly relevant to the ED setting, where a rapid synthesis of information, often within seconds to minutes, informs critical decisions.^17,18^ As a first step in determining whether LLMs might be suitable for this task, it is essential to systematically compare ED-specific LLM-generated one-liners against physician-authored one-liners.

In this study, we evaluated the ability of an LLM to generate ED-specific, one-liner summaries by synthesizing information from clinical notes and imaging reports spanning prior hospital encounters, using an experimental setup that reflects the real-world cognitive demands of ED physicians during patient encounters. To assess the quality of the LLM-generated summaries, we conducted a blinded evaluation in which emergency physicians compared LLM outputs to physician-authored summaries side by side. Each summary was rated along three core dimensions: accuracy, completeness, and clinical utility.

## METHODS

This study was conducted at the University of California, San Francisco (UCSF), in collaboration with the University of California, Berkeley. Patient encounters were drawn from the UCSF Information Commons, a repository of deidentified clinical data and notes certified for research use.^19^ The study protocol was approved by the UC Berkeley Institutional Review Board, and all participants provided written informed consent and were compensated. This study followed the Strengthening the Reporting of Observational Studies in Epidemiology (STROBE) guidelines.^20^

### Cohort Selection

We included ED encounters between March 1, 2022, and March 31, 2024 for patients aged 18 years or older with at least one prior inpatient admission at the UCSF Medical Center. Encounters were included if they contained (1) a discharge summary from an inpatient admission and (2) an ED provider note from a subsequent visit, ensuring longitudinal clinical context for summarization. Encounters were excluded if either (1) or (2) were incomplete, or if the ED note lacked a clear, identifiable one-liner in the History of Present Illness section of the ED note. We prioritized fully documented encounters to enable the evaluation of summary quality under standardized conditions.

To ensure a breadth of clinical presentations were included, we used a two-tiered stratified sampling strategy: rare chief complaints, defined as those occurring in fewer than 10 encounters across the full dataset (e.g., rash, hematuria), were fully retained, while more common presentations (e.g., chest pain, dyspnea, abdominal pain) were proportionally sampled according to their prevalence distribution in the original cohort. Each encounter was randomly assigned to a unique reviewer to avoid duplicating ratings.

### LLM-Based Summary Generation

Using UCSF’s secure, Health Insurance Portability and Accountability Act–compliant Versa Application Programming Interface on Microsoft Azure, we prompted the LLM (OpenAI GPT-4o; temperature = 0.1 to balance determinism and lexical variability; other settings default) to generate ED-specific one-liner summaries in two steps. First, using note-title heuristics (Table S4), we prompted the LLM to choose the notes most relevant to the ED chief complaint. The chief complaint was provided to the model from a structured data field containing free text entered by the triage nurse. Note types that were available for selection included discharge summaries, progress notes, consult notes, history and physical (H&P) notes, imaging reports, echocardiogram reports, and electrocardiogram (ECG) reports. We provided the most recent notes preceding the index ED encounter, which could span multiple prior encounters. The model then selected all notes that it deemed applicable to the chief complaint (up to seven, one note per note type). This first step served two purposes: 1) it substantially reduced computational resources, and 2) it enabled transparency of the model’s behavioral rationale regarding note type selection.

We first had the LLM summarize the selected notes into ED-specific one-liners using zero-shot prompting. However, preliminary outputs revealed substantial temporal confusion, with the model incorrectly attributing previous hospital notes to the index ED encounter. Because this resulted in clinically implausible and disorganized summaries, we ultimately used a few-shot prompting technique. After grouping each case by chief complaint (e.g., chest pain, abdominal pain, altered mental status), we used sentence-transformer representations to embed the discharge summary and chief complaint text. The three most semantically similar cases (k = 3) to the case being summarized were then retrieved as exemplars, each containing a complete physician-authored one-liner. These exemplars were inserted into the prompt as contextual examples, allowing the model to generate a similarly structured, yet specific one-liner for the new case. The full prompt structure, including instruction text, exemplar formatting, and output constraints, is shown in Table S4.

Additionally, pilot testing revealed that the LLM inconsistently used standard medical abbreviations (e.g., “SOB” for shortness of breath, “HTN” for hypertension). This inconsistency not only reduced summary brevity but could also signal to reviewers that the summary was LLM-generated. We therefore addressed this by adding a medical abbreviation guidance document^21^ to the prompt (Table S3).

### One-Liner Summary Evaluation

Twenty-one emergency physicians (12 attendings and 9 residents; Table S1) participated in the blinded evaluation of the one-liner summaries. Each participant reviewed cases using a custom web-based platform (Streamlit) designed to simulate a simplified ED chart interface. For each encounter, reviewers received a standardized packet containing patient demographics (age, sex), chief complaint, and the same most recent notes provided to the LLM (one of each specified type, up to seven total). They were then shown two anonymized one-liner summaries side by side, one written by the original ED physician note writer and one generated by the LLM, presented in randomized order to minimize bias. Reviewers were blinded to which summary was LLM- or physician-authored. Before rating, all participants completed a brief calibration session with detailed instructions covering the evaluation platform and metrics. Reviewers rated each summary using 5-point Likert scales in three domains:

- Accuracy: Is the information factually correct and free of fabrication?
- Completeness: Does it capture all clinically relevant information?
- Clinical utility: How informative is it for decisions in a real-world ED workflow?

Reviewers also indicated a preference between the two summaries (LLM, human, or tie) and could optionally provide brief free-text comments explaining their rationale.

### Statistical Analysis

To evaluate the LLM’s behavioral rationale, we identified the specific note types the model chose to incorporate when generating each one-liner summary. For each one-liner summary, we also calculated the number and proportion of available notes utilized by the LLM. Note selection patterns were then stratified by chief complaint category, with complaints grouped into overarching clinical systems. Descriptive statistics including mean (SD), median, and interquartile range were computed for the number of notes selected overall and by clinical category. To assess whether note selection varied significantly across chief complaint categories, we performed Kruskal-Wallis H-tests. Selection rates for individual note types were calculated as the percentage of encounters where each note type was utilized, both overall and stratified by clinical category, to identify patterns in notes preferences based on clinical presentation.

Reviewer preference data were summarized as win rates: the proportion of encounters in which one summary type was favored over the other. For each summary, the three domain scores were averaged to yield an overall quality score (Appendix S1). Reviewer ratings of summary accuracy, completeness, and clinical utility were analyzed using linear mixed-effects models. Each domain was modeled separately with summary type (LLM- vs physician-authored) as a fixed effect and reviewer as a random intercept to account for clustering of ratings within reviewers. A random intercept for case was initially specified but dropped after contributing negligible variance (i.e., singular fit), as each case was rated by only one reviewer. Model estimates were obtained using restricted maximum likelihood, and p-values were computed via Satterthwaite’s degrees-of-freedom approximation.^39^ Estimated marginal means and 95% confidence intervals were derived for each summary type (LLM- vs Physician-generated) within each evaluation domain. All tests were two-tailed with α = 0.05 significance. Model residuals were inspected to confirm approximate normality and homoscedasticity assumptions. Because each case was evaluated by a single reviewer, interrater reliability could not be estimated; instead, all rating scales were anchored by predefined Likert point descriptors to support scoring consistency across reviewers. We ensured interrater consistency through analysis of the free text justifications collected.

To understand how note characteristics influenced the quality of the one-liner summaries, we first compared physician- and LLM-generated summary performance relative to individual note type lengths. Pearson correlation coefficients were calculated to determine whether note length was associated with quality ratings in either group. Second, we evaluated how specific note types included in the LLM prompt affected the quality of the LLM-generated one-liner summary. For encounters where specific note types were available, we compared the LLM-generated one-liner summary quality between cases where these notes were included in the prompt versus excluded, reporting the difference using Cohen’s d effect size. The note analyses were performed in Python 3.11, and the reviewer ratings analyses were performed in R version 4.3.1.

### Qualitative Analysis

We conducted a rapid content analysis of open-ended survey responses to identify key themes.^22^ Free-text data were reviewed systematically by NG and MM. Using a descriptive analytic approach, we summarized content within each open-ended item and grouped similar responses into thematic categories. This process involved both deductive coding, based on the three evaluation domains, and inductive coding to capture emergent themes raised by participants. A matrix was used to organize and compare responses across participants and questions. Thematic summaries were refined through iterative team discussion, and representative quotes were identified to illustrate major findings.

## RESULTS

### Cohort Characteristics

Of 10,631 ED encounters, 3,984 encounters met the inclusion criteria. A random sample of 300 encounters was selected for evaluation. Each physician reviewed 1-10 non-overlapping cases, resulting in a total of 99 unique ED encounters reviewed with 99 unique patients. Nearly all encounters contained progress notes (99%), H&P notes (98%), and imaging reports (98%). ECGs were present in 87% of encounters, consult notes in 64%, and echocardiogram reports in 56%. The mean (SD) length of all notes combined per case was 5,598 (2,314) words, with 6.0 (range, 2-7) distinct note types included. Note length ranged from 36 (32) words for ECG reports to 2,310 (1,263) words for discharge summaries, with intermediate lengths for imaging (124 [117] words), echocardiograms (677 [109] words), consult notes (854 [629] words), progress notes (917 [1,010] words), and H&P notes (1,332 [770] words)

### LLM-Based Summary Generation

The LLM requested a mean (SD) of 3.73 (0.67) notes (median: 4.0, IQR: 3.0-4.0), utilizing 53.2% (9.5%) of available notes. H&P notes were nearly universally requested (97.0%), followed by imaging reports (81.8%). Consult notes were included in 44.4% of cases, while cardiac-specific notes (ECG: 21.2%, echocardiogram: 9.1%) and progress notes (19.2%) were selectively utilized. Note selection patterns varied significantly by chief complaint category (Kruskal-Wallis H = 17.29, p < 0.001). Cardiopulmonary presentations required the most comprehensive notes (shortness of breath: 4.9(0.3) notes), with universal inclusion of cardiac studies (ECG: 100%, echocardiogram: 88% for shortness of breath). In contrast, musculoskeletal complaints required fewer notes (back pain: 3.2 (0.5) notes) and appropriately excluded cardiac notes. Psychiatric evaluations demonstrated high utilization of progress notes (75%) and consult notes (100%), while avoiding imaging and cardiac studies entirely (Table 1).

**Table 1:** LLM Note Selection Rates by the Top 13 Chief Complaints.

| Category | Chief Complaint | N | Mean | Note Selection Rate (%) |  |  |  |  |  |  |
| --- | --- | --- | --- | --- | --- | --- | --- | --- | --- | --- |
|  |  |  |  | DS | HP | PN | ECG | IM | EC | CN |
| Cardiopulm. | SOB | 8 | 4.9 | 100 | 100 | 0 | 100 | 100 | 88 | 0 |
|  | Chest Pain | 4 | 4.0 | 100 | 75 | 0 | 100 | 100 | 25 | 0 |
| GI | Abd. Pain | 23 | 3.6 | 100 | 96 | 0 | 0 | 100 | 0 | 65 |
| General | Fever | 3 | 3.7 | 100 | 100 | 67 | 0 | 67 | 0 | 33 |
|  | Referral | 3 | 3.0 | 100 | 100 | 67 | 0 | 0 | 0 | 33 |
|  | Fall | 2 | 3.5 | 100 | 100 | 50 | 0 | 100 | 0 | 0 |
| MSK | Back Pain | 4 | 3.2 | 100 | 100 | 0 | 0 | 100 | 0 | 25 |
|  | Foot Pain | 2 | 3.0 | 100 | 100 | 0 | 0 | 100 | 0 | 0 |
| Neuro | Headache | 3 | 3.7 | 100 | 100 | 0 | 0 | 100 | 0 | 67 |
|  | Seizures | 3 | 4.7 | 100 | 100 | 0 | 100 | 100 | 0 | 67 |
|  | Dizziness | 3 | 4.0 | 100 | 100 | 0 | 100 | 100 | 0 | 0 |
|  | Syncope | 2 | 4.0 | 100 | 100 | 0 | 50 | 100 | 50 | 0 |
| Psych | Psych Eval | 4 | 3.8 | 100 | 100 | 75 | 0 | 0 | 0 | 100 |

DS=Discharge Summary; HP=H&P; PN=Progress Notes; IM=Imaging; EC=Echo; CN=Consult
SOB=Shortness of Breath; GI=Gastrointestinal; MSK=Musculoskeletal
Note: Mean represents the average number of notes selected per encounter. N = number of encounters for the given chief complaint.

### One-Liner Summary Evaluation

Across all 99 cases, LLM-generated one-liners were preferred over physician-authored one-liners in 50 cases (50.5%); physician-authored one-liners were preferred in 38 cases (38.4%); 11 cases (11.1%) were rated equally. Excluding ties, the LLM win rate was 56.8% (95% CI, 46.4–66.7%).

In mixed-effects models adjusting for reviewer as a random effect, LLM-generated summaries scored 0.78 points higher for accuracy (95% CI 0.497–1.059; p = 1.98 × 10⁻⁷), 0.43 points higher for completeness (95% CI 0.152–0.716; p = 0.0030), and 0.67 points higher for clinical utility (95% CI 0.360–0.973; p = 3.28 × 10⁻⁵). The overall composite score similarly favored the LLM (β = 0.626, SE = 0.131; 95% CI 0.369–0.884; p = 3.82 × 10⁻⁶). Model-estimated marginal means (± 95% CI) were 4.18 (3.83–4.54) vs 3.40 (3.05–3.76) for accuracy, 3.69 (3.37–4.00) vs 3.25 (2.94–3.57) for completeness, and 3.88 (3.47–4.28) vs 3.21 (2.81–3.61) for clinical utility, for LLM-generated and physician-authored summaries respectively. Case-level variance was estimated at zero (reflecting the single-reviewer-per-case design), while reviewer-level variance was non-zero (SD ≈ 0.65–0.80), indicating individual differences in rating stringency. Overall, LLM-generated summaries were judged by physician reviewers as more accurate, complete, and clinically useful.

### Source Note Characteristics and Summary Performance

#### Impact of Note Length between Physician vs LLM

For LLM-generated summaries, overall note length was not associated with accuracy (r = 0.053, p > 0.05), completeness (r = 0.039, p > 0.05), or clinical utility (r = 0.094, p > 0.05). In contrast, physician-authored summary completeness increased with longer notes (r = 0.229, p = 0.023). Stratifying our analysis by specific note type revealed that for LLM-generated summaries, longer history and physical notes were associated with higher accuracy (r = 0.243, p = 0.016), whereas longer progress notes were negatively correlated with completeness (r = 0.229, p = 0.023) and overall quality (r = 0.210, p = 0.038).

#### Impact of Selective Note Inclusion on LLM-Generated Summaries

The effect of selective note inclusion by the LLM was also evaluated. When imaging reports were both available and selected by the LLM, summary quality scores were higher (mean 4.05, SD 0.93) compared with encounters where imaging reports were available but not included (mean 3.58, SD 1.14; Cohen’s d = 0.49; p = 0.078).

Figure 2A shows correlations between individual note type lengths and LLM-generated summary quality across domains. Figure 2B displays the effect of explicitly including individual note types in the LLM input. Imaging reports (Cohen’s d = 0.42) and echocardiograms (Cohen’s d = 0.32) were associated with improved summary quality compared to encounters where these notes were available but not included, while inclusion of progress notes had a small negative effect.

**Figure 1:**
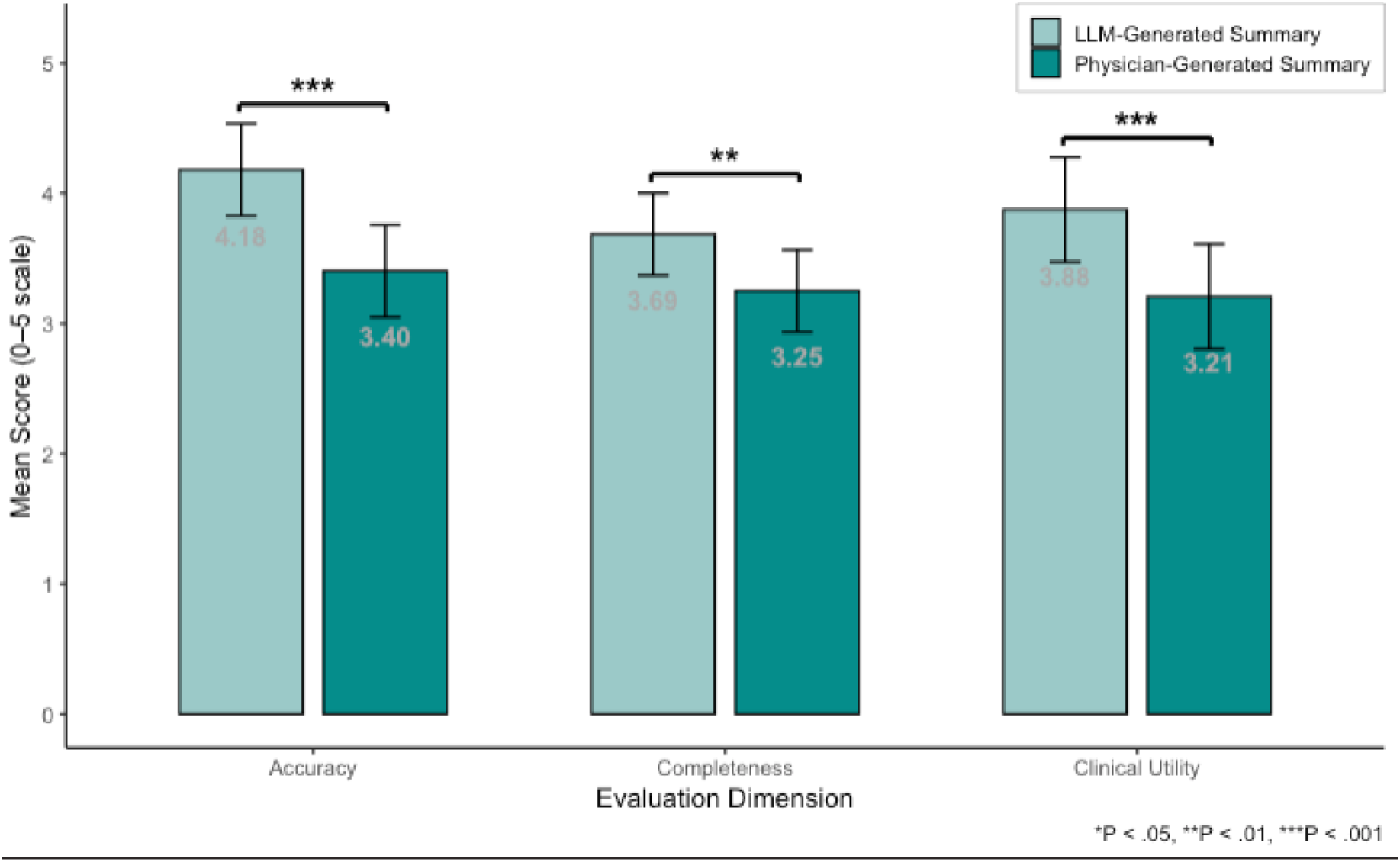
Comparison of LLM-generated and physician-authored one-liner summaries across evaluation dimensions. Mean scores (± 95% CI) for LLM-generated summaries (light teal bars) and physician-authored summaries (dark teal bars) on a 5-point Likert scale across accuracy, completeness, and clinical utility. Significance was based on the model-estimated confidence interval of the paired mean difference.

**Figure 2:**
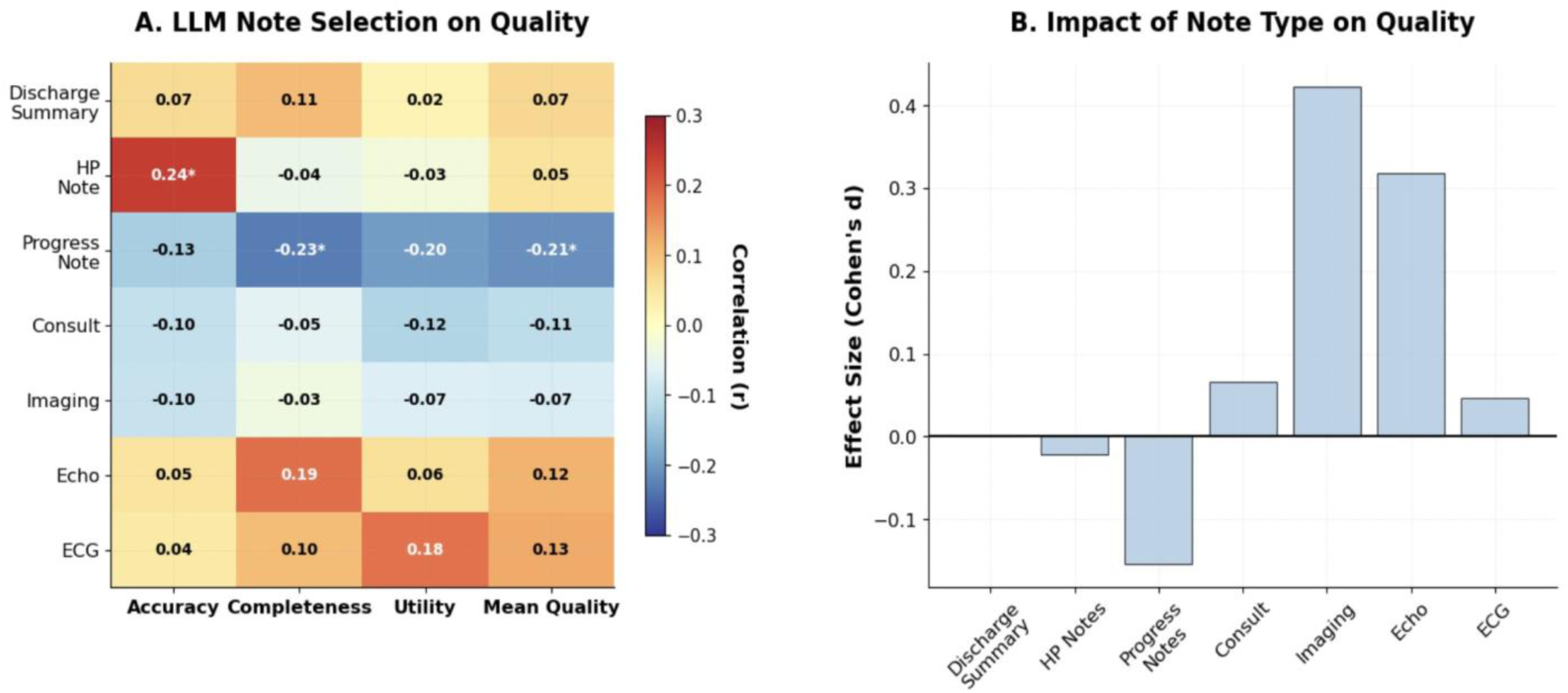
Impact of note characteristics on LLM summary quality. (A) Correlations (*r*) between individual note type lengths (decreasing in length from top to bottom of the matrix) and quality scores across domains (accuracy, completeness, clinical utility, and mean quality). (B) Effect sizes (Cohen’s *d*) showing the impact of explicitly including specific note types on LLM-generated summary quality compared to when those note types were available but not included. Asterisks indicate statistically significant correlations (p < 0.05).

### Qualitative Analysis

Fifty-five paired evaluations had free-text commentary in which reviewers explained their preferences between LLM- and physician-authored summaries. The major themes emerging from these explanations are shown in Table 2.

**Table 2.** Themes from Reviewer Free-Text Explanations of One-Liner Preferences.

| Theme | Representative Quotes |
| --- | --- |
| Importance of factual reliability as measured by alignment with source documentation | “Does not have invented PMH; accuracy matters even if incomplete.” *(preferred LLM)*<br>“She’s not actually on oxygen which is why I don’t like A.” *(preferred LLM)*<br>“Both of them indicate he had transverse sinus thrombosis, which it didn’t actually appear to have on final MRI (though there was initial concern for it).” *(equal preference)* |
| Inclusion of clinical context relevant to presenting complaint can help guide differential diagnosis | “History of MDR bacteriuria relevant to abdominal pain.” *(preferred Physician)*<br>“Colors our approach to this person’s abdominal pain.” *(preferred Physician)*<br>“Mentions chest pain with abdominal pain, changing DDX.” *(preferred Physician)*<br>“Misses critically important information about the patient’s history with arrhythmia, which could lead to a dizziness that may have preceded the fall.” *(preferred LLM)*<br>“Includes residual stroke deficits relevant to the presentation.” *(preferred LLM)* |
| Accurate references to recency and timing of key clinical events are essential | “Gives the specifics of the procedure in a very concise way and actually gets the date right.” *(preferred LLM)*<br>“Many of the problems in the one-liner were from a recent hospitalization (as opposed to not knowing how recent they were).” *(preferred LLM)* |
| Added value of actionable information supporting triage or management decisions | “Would prompt me to see the patient sooner since it mentioned unstable vitals.” *(preferred Physician)*<br>“Includes who to involve as consultants.” *(preferred Physician)*<br>“Includes pertinent information about the specific form of cancer, location of mets, and a recent admission for sepsis which significantly changes my initial approach to this pt” *(preferred LLM)* |

During the content analysis, five major themes emerged: (1) Importance of factual reliability as measured by alignment with source documentation, (2) Inclusion of relevant clinical context to presenting complaint can help guide differential diagnosis, (3) Accurate references to recency and timing of key clinical events are essential, and (4) Added value of actionable information supporting triage or management decisions (Table 2).

Reviewers often praised LLM-generated summaries for their factual reliability and adherence to charted information. Physician-authored summaries were favored when they had richer contextual framing, including key comorbidities and diagnostic nuances (“History of MDR bacteriuria, relevant to abdominal pain”) that prompted actionable items. Reviewers especially valued summaries that guided clinical prioritization or next steps, for example, noting key contextual details (e.g., variceal banding) that informed which consultants to involve early. In some cases, reviewers found the LLM and physician summaries equally useful but non-overlapping, highlighting complementary strengths and suggesting that combining them could yield more clinically salient, useful summaries: “Both statements contain something that the other is missing which would be helpful (ex: B leaves out the fact patient is not taking AC [anticoagulation] for known DVT [deep venous thrombosis], A leaves out what specific substance, etc.)”.

## DISCUSSION

In this study, we found that an LLM, when prompted with domain-specific context, generated one-liner summaries that not only significantly outperformed physician-authored counterparts across accuracy, completeness, and clinical utility, but were also preferred in slightly over half of cases. Secondary analysis showed that the LLM’s selective note-inclusion strategy, using only about half of the available documents and drawing more heavily from structured or diagnostically relevant notes, was associated with higher summary quality. While reviewers described physician-authored summaries as offering contextual nuance, LLM-generated summaries were described as more factually aligned with the chart, temporally accurate, and inclusive of key details.

A growing body of work has examined LLMs in clinical summarization, but nearly all prior studies have focused on long-form documentation—such as discharge summaries or inpatient visit notes—rather than the concise summaries used in emergency care.^11–13, 23^ Prior literature has also relied almost exclusively on single-encounter inputs, typically structured notes, and has not examined whether LLMs can integrate information spanning multiple prior encounters, heterogeneous data types (e.g., clinical notes, imaging, ECG reports), and temporally distant clinical events into a single actionable statement.^14^ Our study therefore adds to the literature by demonstrating that an LLM can, with few-shot prompting, integrate information across encounters and data sources to produce an accurate, complete, and useful one-liner summary. Importantly, we allowed the model to select its own source documents, mirroring clinical information triage and avoiding degradation from indiscriminate long-input ingestion. Therefore, we extend the clinical LLM-assisted summarization literature by showing how prompting strategy, input structure, and selective note inclusion impact summary quality.

Our study demonstrated that LLM performance critically depended on how context was structured during input. In zero-shot settings, the model frequently collapsed timelines, misrepresenting chronic conditions as acute complaints or misplacing prior diagnostic test results, errors with potentially serious clinical consequences.

These patterns reflect well-documented failures in temporal grounding for EHR summarization. ^23^ The kNN few-shot prompting technique improved temporal coherence and clinical specificity, consistent with previous work on domain-specific prompting and retrieval augmented generation in clinical NLP.^24^ This highlights that high-quality ED summarization across multiple prior healthcare encounters requires not only structured context and curated exemplars but also an explicit consideration of time to avoid clinically dangerous distortions.

Our findings also underscore the importance of careful curation of note types and lengths for operational deployment. The model did not simply ingest all available documentation; rather, it demonstrated context-sensitive document selection, incorporating only ∼53% of notes while maintaining clinical appropriateness. This selective behavior mirrors expert physician heuristics^25–27^ and varied consistently with chief complaint; for example, cardiac documents were nearly always selected for cardiopulmonary presentations and rarely for musculoskeletal or psychiatric cases. This “clinical mimicry”^28^ is not only important for accuracy, but also for efficiency and broader implementation in health systems, where token costs accumulate rapidly; any real-world deployment will require selective note strategies to remain operationally viable.^29^ Our note-level analyses showed that high-yield documents (e.g., H&P notes, imaging reports) contributed disproportionately to summary quality, whereas verbose progress notes diluted signal. These effects may reflect both length-related dilution of relevant signal and recency or “primary source” bias, where more recent or central documents disproportionately influence summarization.^35^ Together, these findings highlight that targeted retrieval of concise, clinically salient documents, rather than indiscriminate ingestion of all available text, is essential for reliable, cost-effective deployment of generative summarization in the ED.

Despite the context-sensitive note selection demonstrated by our LLM, contemporary LLMs (e.g., GPT-4o) offer little to no visibility into how they choose and prioritize clinical documents,^30^ muddling transparency for ED physicians who must quickly verify the source and accuracy of critical information. The reduced cognitive load and improved operational efficiency of automated one-liner generation can only be realized if ED physicians are provided with mechanisms to rapidly validate model-derived content, as ED physicians lack the time to cross-check autogenerated summaries against the full chart. Our note-selection and qualitative analyses suggest a practical solution: enable the model to surface excerpt-level citations or highlight specific note types underlying each piece of information. Such source transparency would allow clinicians to quickly verify key details without navigating long documents, balancing efficiency with oversight. As health systems consider deploying LLM-assisted chart summarization, incorporating a transparent attribution mechanism will be essential to establish clinician trust and safe adoption in time-pressured environments.

The complementary error profiles between LLM- versus physician-authored summaries highlighted in the qualitative analyses suggest that an LLM may be used to draft the core clinical history, while physicians can add contextual nuance. We found that the LLM’s performance remained stable regardless of note volume, suggesting robustness to information density and fragmentation—conditions known to contribute to cognitive overload and fatigue in ED physicians.^33,34^ This stability under high-volume input is especially advantageous in the ED environment, which requires a longitudinal synthesis of clinical information. Developing a hybrid workflow would therefore play to the strengths of both the LLM and the physician: the LLM could ensure completeness and neutral phrasing, while the physician could review and contextualize the information relevant to the clinical presentation.^32^

Lastly, our findings carry important implications for the workflow of emergency physicians. An LLM that automatically produces ED-specific one-liner summaries could potentially reduce the time spent on chart review, support rapid cognitive framing during high-acuity encounters, enhance the completeness of handoffs, and ease documentation burden. By front-loading information synthesis, LLMs would enable clinicians to focus on higher-order reasoning and patient interaction. Collectively, these improvements have the potential to lessen burnout, which remains pervasive in emergency medicine.^6,7, 33, 36–38^ While our retrospective study provides the first step toward achieving these outcomes, prospective validation will ultimately be requires. However, implementation must prioritize transparency and safety; these tools should augment, not replace, clinical reasoning.^9^

## LIMITATIONS

First, physician-authored one-liners may have incorporated clinically relevant context that was not present in the extracted notes provided to reviewers—most commonly nuances surrounding the chief complaint, which originated from a structured field rather than narrative documentation. Reviewers occasionally penalized physician summaries when these verbally obtained details were not present in the excerpted notes, which may have led to slightly inflated ratings in favor of LLM-generated summaries. However, our qualitative review of free-text comments indicated that such instances were relatively rare and unlikely to meaningfully alter our findings. Second, the cohort was drawn from a single academic center, limiting generalizability. However, our quaternary academic ED provides care for highly complex patients with extensive longitudinal histories and multidisciplinary involvement, suggesting that performance may be equal or better in less specialized settings. Third, our evaluation was retrospective and did not assess downstream impact, which was beyond the scope of our study. Finally, although we did not test other prompting or fine-tuning strategies, these would likely reinforce or improve the performance demonstrated here, given that our results were achieved with minimal optimization of an out-of-the-box model.

## CONCLUSIONS

In this blinded evaluation, LLM-generated one-liner summaries achieved higher scores for accuracy, completeness, and clinical utility, and were preferred over physician-authored summaries. The model’s selective note inclusion, mirroring physician heuristics, suggests potential for efficiency in real-world deployment while maintaining clinical appropriateness. These findings suggest that, under structured prompting and clinical oversight, generative models can be used to synthesize temporally fragmented notes into coherent and clinically useful summaries that support ED decision-making.^10,11^

## Data Availability

Data may be made available upon request at the discretion of the study authors.

## Acknowledgements

We thank the participating emergency physicians for their time and expertise in evaluating the summaries.

Versa (UCSF’s secure ChatGPT 4o by Open AI) was used on August 10, 2025 to revise grammatical errors and sentences for clarity. Dr. Molina takes responsibility for the integrity of the content generated.

## REFERENCES

1. Yadav S, Rawal G, Jeyaraman M. Decision Fatigue in Emergency Medicine: An Exploration of Its Validity. Cureus. 2023;15(12):e51267. Published 2023 Dec 29. doi:10.7759/cureus.51267

2. Moskop JC, Sklar DP, Geiderman JM, Schears RM, Bookman KJ. Emergency department crowding, part 1--concept, causes, and moral consequences. Ann Emerg Med. 2009;53(5):605–611. doi:10.1016/j.annemergmed.2008.09.019

3. Heilman JA, Flanigan M, Nelson A, Johnson T, Yarris LM. Adapting the I-PASS handoff program for emergency department inter-shift handoffs. West J Emerg Med. 2016;17(6):756–761. doi:10.5811/westjem.2016.9.30574

4. Greenwald JL, Denham CR, Jack BW. The hospital discharge: a review of a high risk care transition with highlights of a reengineered discharge process. J Patient Saf. 2007;3(2):97–106.

5. Pivovarov R, Elhadad N. Automated methods for the summarization of electronic health records. J Am Med Inform Assoc. 2015;22(5):938–947. doi:10.1093/jamia/ocv032

6. Laxmisan A, Hakimzada F, Sayan OR, Green RA, Zhang J, Patel VL. The multitasking clinician: decision-making and cognitive demand during and after team handoffs in emergency care. Int J Med Inform. 2007;76(11-12):801–811. doi:10.1016/j.ijmedinf.2006.09.019

7. Moy AJ, Hobensack M, Marshall K, et al. Understanding the perceived role of electronic health records and workflow fragmentation on clinician documentation burden in emergency departments. J Am Med Inform Assoc. 2023;30(5):797–808. doi:10.1093/jamia/ocad038

8. Peccoralo LA, Kaplan CA, Pietrzak RH, Charney DS, Ripp JA.The impact of time spent on the electronic health record after work and of clerical work on burnout among clinical faculty. J Am Med Inform Assoc. 2021;28(6):1016–1026. doi:10.1093/jamia/ocaa349

9. ALQahtani DA, Rotgans JI, Mamede S, et al. Does Time Pressure Have a Negative Effect on Diagnostic Accuracy?. Acad Med. 2016;91(5):710–716. doi:10.1097/ACM.0000000000001098

10. Singhal K, Azizi S, Tu T, et al. Large language models encode clinical knowledge. Nature. 2023;620(7974):172–180. doi:10.1038/s41586-023-06291-2

11. Van Veen D, Van Uden C, Blankemeier L, et al. Adapted large language models can outperform medical experts in clinical text summarization. Nat Med 30, 1134–1142 (2024). 10.1038/s41591-024-02855-5

12. Aali A, van Veen D, Arefeen YI, et al. A dataset and benchmark for hospital course summarization with adapted large language models. J Am Med Inform Assoc. 2025;32(3):470–479. doi:10.1093/jamia/ocae312

13. Williams CYK, Scheinker D, Samore M, et al. Evaluating large language models for drafting emergency department discharge summaries. medRxiv. Preprint posted online December 14, 2023. doi:10.1101/2023.12.13.23299561

14. Bednarczyk L, Croxford E, Kannan V, et al. Scientific evidence for clinical text summarization using large language models: scoping review. J Med Internet Res. 2025;27:e68998 doi: 10.2196/68998

15. Johnson AEW, Pollard TJ, Shen L, et al. MIMIC-III, a freely accessible critical care database. Sci Data. 2016;3:160035.

16. Johnson AEW, Bulgarelli L, Pollard TJ, et al. MIMIC-IV: a freely accessible electronic health record dataset. Sci Data. 2023;10(1):113.

17. Menachemi N, Collum TH. Benefits and drawbacks of electronic health record systems. Risk Manag Healthc Policy. 2011;4:47–55. doi:10.2147/RMHP.S12985

18. Jiang S, Shen S, Agrawal M, et al. Conceptualizing machine learning for dynamic information retrieval of electronic health record notes. arXiv. Preprint posted online 2023. doi:10.48550/arXiv.2306.XXXX

19. Radhakrishnan L, Schenk G, Muenzen K, et al. A certified de-identification system for all clinical text documents for information extraction at scale. JAMIA Open. 2023;6(3):ooad045. doi:10.1093/jamiaopen/ooad045

20. von Elm E, Altman DG, Egger M, Pocock SJ, Gøtzsche PC, Vandenbroucke JP. The Strengthening the Reporting of Observational Studies in Epidemiology (STROBE) statement: guidelines for reporting observational studies. Lancet. 2007;370(9596):1453–1457.

21. American Speech-Language-Hearing Association. Common medical abbreviations. Accessed July 23, 2025. https://www.asha.org/practice-portal/professional-issues/documentation-in-health-care/common-medical-abbreviations/

22. Nevedal AL, Reardon CM, Opra Widerquist MA, et al. Rapid versus traditional qualitative analysis using the consolidated framework for implementation research (CFIR). Implement Sci. 2021;16(1):67. doi:10.1186/s13012-021-01111-5

23. Verma R, Gopinath D, Chen RJ, et al. Verifiable summarization of EHRs using LLMs to support chart review. medRxiv. Preprint posted online April 4, 2025. doi:10.1101/2025.04.02.24130984

24. Sivarajkumar S, Kelley M, Samolyk-Mazzanti A, Visweswaran S, Wang Y. An Empirical Evaluation of Prompting Strategies for Large Language Models in Zero-Shot Clinical Natural Language Processing: Algorithm Development and Validation Study. JMIR Med Inform. 2024;12:e55318 doi: 10.2196/55318

25. Rajkomar, A., Oren, E., Chen, K. et al. Scalable and accurate deep learning for electronic health records. NPJ Digit Med. 2018;1:18. 10.1038/s41746-018-0029-1

26. Pelaccia T, Tardif J, Triby E, et al. How and when do expert emergency physicians generate and evaluate diagnostic hypotheses? A qualitative study using head-mounted video cued-recall interviews. Ann Emerg Med. 2014;64(6):575–585. doi:10.1016/j.annemergmed.2014.05.003

27. Kovacs G, Croskerry P. Clinical decision making: an emergency medicine perspective. Acad Emerg Med. 1999;6:947–952.

28. Wang T, Yang C, Zou M, et al. A study of extractive summarization of long documents incorporating local topic and hierarchical information. Sci Rep. 2024;14(1):10140. Published 2024 May 2. doi:10.1038/s41598-024-60779-z

29. Grote T, Berens P. On the ethics of algorithmic decision-making in healthcare. J Med Ethics. 2020;46(3):205–211.

30. Amann J, Blasimme A, Vayena E, Dietmar F, Madai V. Explainability for artificial intelligence in healthcare. BMC Med Inform Decis Mak. 2020;20(1):173.

31. Ghassemi M, Oakden-Rayner L, Beam AL. The false hope of current approaches to explainable artificial intelligence in health care. Lancet Digit Health. 2021;3(11):e745–e750. doi:10.1016/S2589-7500(21)00208-9.

32. Lai, Y., Kankanhalli, A., & Ong, D. (2021). Human-AI Collaboration in Healthcare: A Review and Research Agenda. In Proceedings of the 54th Hawaii International Conference on System Sciences (HICSS) (pp. 4663–4672). IEEE. 10.24251/HICSS.2021.046

33. Asgari E, Kaur J, Nuredini G, et al. Impact of Electronic Health Record Use on Cognitive Load and Burnout Among Clinicians: Narrative Review. JMIR Med Inform. 2024;12:e55499. Published 2024 Apr 12. doi:10.2196/55499

34. Kim D, Preiksaitis C, Rose C. Feasibility of real-time cognitive load assessment using physiologic signals in the emergency department. medRxiv. doi: 10.1101/2025.06.02.25328813

35. Guo X, Vosoughi S. Serial position effects of large language models. arXiv. Preprint posted online June 25, 2024. https://arxiv.org/abs/2406.15981

36. Arndt BG, Beasley JW, Watkinson MD, et al. Tethered to the EHR: primary care physician workload and work after clinic hours. Ann Fam Med. 2017;15(5):419–426.

37. Sinsky C, Colligan L, Li L, et al. Allocation of physician time in ambulatory practice: a time and motion study in 4 specialties. Ann Intern Med. 2016;165(11):753–760.

38. Alobayli F, O’Connor S, Holloway A, Cresswell K. Electronic Health Record Stress and Burnout Among Clinicians in Hospital Settings: A Systematic Review. Digit Health. 2023;9:20552076231220241. Published 2023 Dec 19. doi:10.1177/2055207623122024

39. Kuznetsova, A., Brockhoff, P. B., & Christensen, R. H. B. (2017). lmerTest Package: Tests in Linear Mixed Effects Models. Journal of Statistical Software, 82(13), 1–26. 10.18637/jss.v082.i13

