## Supplement 1 for "Physician- versus Large Language Model-Generated Clinical Summaries in the Emergency Department"

**Supplementary Material**

| Table of Contents | Page Number |
| --- | --- |
| 1. Table S1. Physician Reviewer Characteristics | 1-3 |
| 2. Table S2. Patient Encounter Characteristics | 4-5 |
| 3. Appendix S1. kNN Few-Shot Summarization With Dynamic Note Selection (Method Summary) | 6-7 |
| 4. Table S3. Retrieval and Generation Hyperparameters | 8 |
| 5. Table S4. Prompt Templates for Note Selection and One-Liner Generation | 9-10 |
| 6. Appendix S2. Instructions Provided to Physician Participants | 11-13 |
| 7. Figure S2. Study Methodology and LLM Pipeline | 14 |

### Table S1. Physician Reviewer Characteristics

| **Characteristic** | **Value** |
| --- | --- |
| **Basic Demographics**  Age, mean (SD), y | 36.3 (9) |
| Age, median (IQR), y | 32.0 (30.0–43.0) |
| Age range, y | 27–55 |
| Sex, No. (%)  Male | 11 (52.3) |
| Female | 8 (38.1) |
| Non-binary | 1 (4.8) |
| Prefer not to say | 1 (4.8) |
| **Professional Characteristics**  Credential type, No. (%)  Attending | 12 (57.14) |
| Resident | 9 (42.84) |
| ER experience, mean (SD), y | 6.8 (6.0) |
| ER experience range, y | 2–23 |
| **AI Experience and Attitudes**  Clinical AI experience level, No. (%)  Minimal | 9 (42.3) |
| Extensive | 4 (19.0) |
| None | 6 (28.6) |
| Moderate | 2 (9.5) |
| Clinical AI openness, No. (%)  Very open | 11 (52.4) |
| Somewhat open | 5 (23.8) |
| Neutral | 4 (19.0) |
| Somewhat resistant | 1 (4.8) |
| AI tools used (Select all that applies), No. (%)  OpenEvidence | 4 (15.4) |
| Perplexity | 2 (7.7) |
| ChatGPT | 2 (7.7) |
| Other AI tools | 1 (3.8) |
| Epic-integrated AI (e.g., Dragon) | 1 (3.8) |
| Claude | 1 (3.8) |
| Gemini/Bard | 1 (3.8) |

### Table S2. Patient Encounter Characteristics

| **Characteristic** | **Value** |
| --- | --- |
| Age, mean (SD), y | 57.0 (21.0) |
| Age, median (IQR), y | 62.0 (37.0–74.5) |
| Age range, y  Age group, No. (%) | 18–88 |
| 65+ | 45(45.5) |
| 45–64 | 23(23.2) |
| 30–44 | 19(19.2) |
| 19–29 | 8(8.1) |
| =18  Sex, No. (%) | 4(4.0) |
| Female | 55(55.6) |
| Male  Race/ethnicity (top 5), No. (%) | 44(44.4) |
| White | 39(39.4) |
| Asian | 26(26.3) |
| Other | 15(15.2) |
| Black or African American | 14(14.1) |
| Native Hawaiian or Other Pacific Islander | 2(2.0) |
| Other race (2 categories)  **Clinical Characteristics**  Top 10 chief complaints, No. (%) | 3(3.0) |
| Abdominal pain | 23(23.2) |
| Shortness of breath | 8(8.1) |
| Psychiatric evaluation | 4(4.0) |
| Back pain | 4(4.0) |
| Chest pain | 4(4.0) |
| Referral | 3(3.0) |
| Headache | 3(3.0) |
| Seizures | 3(3.0) |
| Fever | 3(3.0) |
| Dizziness | 3(3.0) |
| Total unique chief complaints  Top 10 ED diagnoses, No. (%) | 48 |
| Chest pain, unspecified type  Acute Abdominal Pain  Acute nonintractable headache, unspecified  Seizure  Abdominal Pain, epigastric  Sepsis, unspecified organism  Abdominal Pain, left upper quadrant  Fever, unspecified cause  Suiciudal ideation  Pneumonia, unspecified organism  Total Unique ED Diagnoses | (5(5.1)  4(4.0)  4(4.0)  3(3.0)  2(2.0)  2(2.0)  2(2.0)  2(2.0)  2(2.0)  2(2.0)  80 |

### Appendix S1. kNN Few-Shot Summarization With Dynamic Note Selection (Method Summary)

**Overview** We implement a two–stage pipeline that (1) retrieves *k* clinically similar, high– quality one–liner examples via dual–source embeddings and (2) prompts an LLM to *first* select which note types to review (dynamic input curation) and *then* generate a temporally coherent ED one–liner with abbreviation guidance and few–shot exemplars.

#### Stepwise Procedure

1. **Candidate pool:** Start with ED encounters that contain (a) a chief complaint (CC), (b) prior EHR notes (e.g., discharge summary, progress, H&P, imaging, ECG, echo, consult), and (c) a human one–liner (for exemplars).
2. **Embeddings:** Compute Bio ClinicalBERT embeddings for (i) the current case CC and (ii) the current discharge summary (if present). Do the same for all candidate exemplars.
3. **Similarity:** Compute cosine similarities for CC and discharge summary separately; combine with equal weights (0.5/0.5) to yield a single similarity score per candidate.
4. **kNN selection:** Select the top *k* = 3 most similar cases with ground–truth one–liners to use as few–shot exemplars (fall back to fewer if needed).
5. **Dynamic note selection:** Present the LLM with the list of *available* note types for the current case and ask it to return a *comma–separated list* of the notes it wants to read, guided by the chief complaint (hard rule: always include discharge summary if available). Filter out unavailable notes.
6. **Input assembly:** For each requested note, extract content (with date), and concatenate into the *PAST Medical Records* section.
7. **Few–shot context:** Append the *k* exemplar pairs (Example Chief Complaint, Expected Summary) to the prompt.
8. **Abbreviation guidance:** Provide a compact list mapping terms to standard ED abbreviations to enforce concise, specialty–consistent phrasing.
9. **Generation:** Prompt the model with a strict temporal framing: summarize *past* history and *end* with the *current* chief complaint (unaltered), avoiding inference about present illness.
10. **Operational safeguards:** Batch processing (default size 10), GPU use for embeddings, retry logic for API calls (up to 3), caching of embeddings, and default fallbacks if the selection step fails (e.g., at least Discharge Summary).

**Rationale** Dual–source embeddings (CC + Discharge Summary) improve case retrieval relevance; dynamic note selection constrains input volume and focuses on clinically high– yield sources, reducing token cost and improving scalability for health–system deployment. Abbreviation guidance enforces ED style, while few–shot exemplars improve temporal coherence and structure.

### Table S3. Retrieval and Generation Hyperparameters

| **Component** | **Setting / Value** |
| --- | --- |
| Embedding model | Bio ClinicalBERT (CLS embedding) |
| Similarity metric | Cosine similarity |
| Similarity fusion | CC (0.5) + Discharge Summary (0.5) |
| *k* (nearest neighbors) | 3 (fallback to available) |
| Batch size | 10 |
| LLM temperature | 0.1 |
| LLM max tokens (generation) | 4096 |
| Selection retries / Gen retries | up to 3 / up to 3 |
| Abbreviation guidance | Included (domain list) |
| Few–shot exemplars | Included (CC + human one–liner) |
| Defaults on selection failure | Discharge Summary |

| **Prompt** | **Role** | **Content (verbatim)** |
| --- | --- | --- |
| **Note Selection** | System | You are an experienced emergency department (ED) who is preparing to write your note for a patient ED visit. Your task is to decide which medical notes you want to read to create the most accurate and comprehensive summary for your one-liner in your note. You should always start with the discharge summary. |

### Table S4. Prompt Templates (Selection and Generation)

| **Note Selection** | User | Patient basic info: {age}yo {sex} with chief complaint: {chief complaint} Available notes (respond ONLY with the names of notes you want to see, separated by commas): - Discharge  Summary: {Available/Not available} Progress Notes: {Available/Not available}  - H&P: {Available/Not available} -  Echo: {Available/Not available} Imaging: {Available/Not available} Consult: {Available/Not available} - ECG:  {Available/Not available}  Based on the chief complaint, list  ONLY the note types you need to review (comma-separated, no explanation): |
| --- | --- | --- |
| **One–Liner**  **Generation** | System | You are an experienced emergency department  (ED) physician creating a one-liner for a  NEW patient who has just arrived at the ED. The patient’s past medical records are available to you. Your task is to summarize the patient’s relevant PAST medical history and end with their CURRENT chief complaint that is given with no adjectives about the chief complaint as you can NOT assume anything about their current condition. All notes and medical records provided are from PAST encounters, not the current visit. |

| **One–Liner**  **Generation** | User | Create a concise one-liner summary for a patient who has just arrived at the Emergency Department on {arrival date}. The one-liner must:  1. Start with demographic information (age, sex) 2. Include a concise summary of relevant PAST medical history from previous visits/notes 3. End with just CURRENT presenting chief complaint that is not capitilized in the summary and does have additional information regarding the chief complaint: ’{chief complaint}’  IMPORTANT: Everything in the notes is from PAST encounters. The patient is NOW presenting with a NEW complaint:  ’{chief complaint}’.  Use medical abbreviations where appropiate: |
| --- | --- | --- |

{abbreviationguidance}

Here are some example summaries for similar cases: {fewshot examples}

Now, create a similar summary for this new case: Current Chief Complaint: {chief complaint} Age: {age} Sex: {sex} Current ED Arrival Date: {arrival date}

PAST Medical Records: {notes content}

### Appendix S2. Instructions Provided to Physician Participants

Prior to initiating the evaluation, participants were shown the following on-screen instructions in the Streamlit-based application used for the study.

### Study Overview

You will be reviewing patient cases from the emergency department. For each case, you’ll complete a summarization evaluation task.

### Patient Record Review

For each case, you will first review the patient’s medical record, which includes:

- Current chief complaint
- Relevant clinical documentation (e.g., ECG, echocardiogram) typically available during ED workflow

You may review the patient’s record by clicking on the appropriate tabs at the top of the screen or by using the sidebar while completing the evaluation task. Please take your time to review the materials thoroughly before proceeding.

### Summarization Task

After reviewing the patient record, you will evaluate two different one-liner summaries of the case. These are intended to represent the type of one-liner you would write as the ED provider during the patient’s current visit.

You will rate each summary on three dimensions:

1. **Accuracy**: Is the information in the summary factually correct (i.e., free from false or fabricated content)?

**Table S5: Accuracy Evaluation Criteria**

| **Score** | **Description** |
| --- | --- |
| **5** | Completely accurate; no errors or hallucinated information. |
| **4** | Mostly accurate; minor errors unlikely to impact care. |
| **3** | Partially accurate; includes some fabricated content that may affect care. |
| **2** | Mostly inaccurate; significant falsehoods impacting care. |
| **1** | Dangerous inaccuracies; could result in patient harm. |

2. **Completeness**: Does the summary capture all critical and relevant historical or contextual information?

**Table S6: Completeness Evaluation Criteria**

| **Score** | **Description** |
| --- | --- |
| **5** | All critical information required for clinical decision-making is included. |
| **4** | Most important elements included; minor omissions present. |
| **3** | Basic information present; several key elements are missing. |
| **2** | Major omissions of important clinical data. |
| **1** | Critically incomplete; unsuitable for clinical use. |

3. **Clinical Utility**: Would this summary be helpful in a real-world clinical workflow?

After evaluating both summaries, you will select which one is better overall and provide a brief explanation for your choice.

**Table S7: Clinical Utility Evaluation Criteria**

| **Score** | **Description** |
| --- | --- |
| **5** | Would significantly improve clinical workflow and save time. |
| **4** | Would somewhat improve workflow. |
| **3** | Neutral impact on workflow. |
| **2** | Would somewhat impede workflow. |
| **1** | Would significantly impede workflow or increase workload. |

### Navigation and Progress

You may navigate between patient encounters using the buttons at the bottom of each page. Your progress is saved automatically upon submission of each evaluation.

*Note: An instructional image (see below) was also displayed in the interface. Participants could not proceed without viewing this screen.*

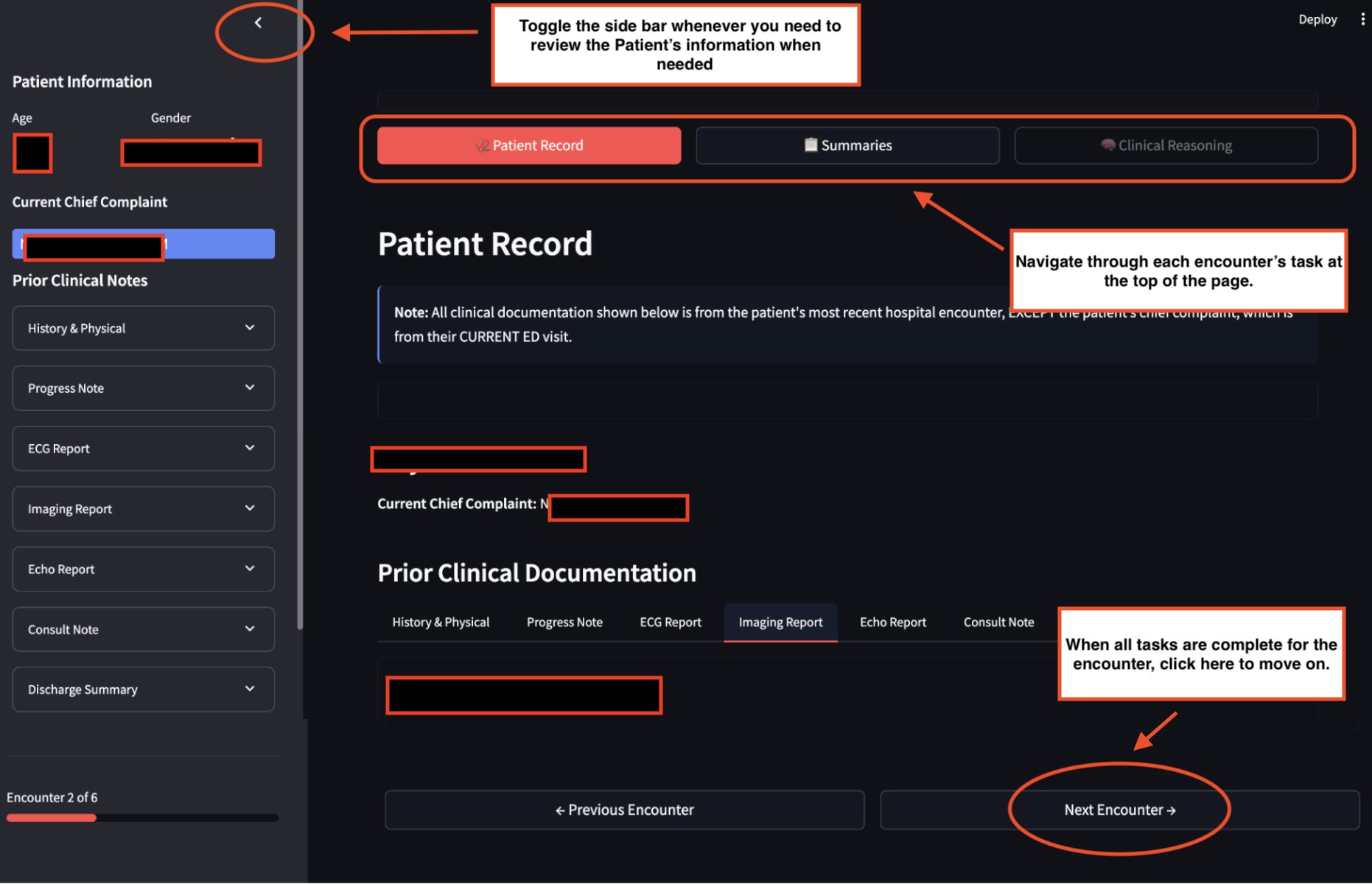

**Figure S1: Instructional Image with Redacted Patient-Sensitive Information**

### Figure S2. Study Methodology and LLM Pipeline: (A) systematic cohort selection from electronic health records using inclusion criteria and stratified sampling, and (B) LLM-based pipeline for generating one-liner clinical summaries from multisource clinical data, with K-nearest neighbors approach.

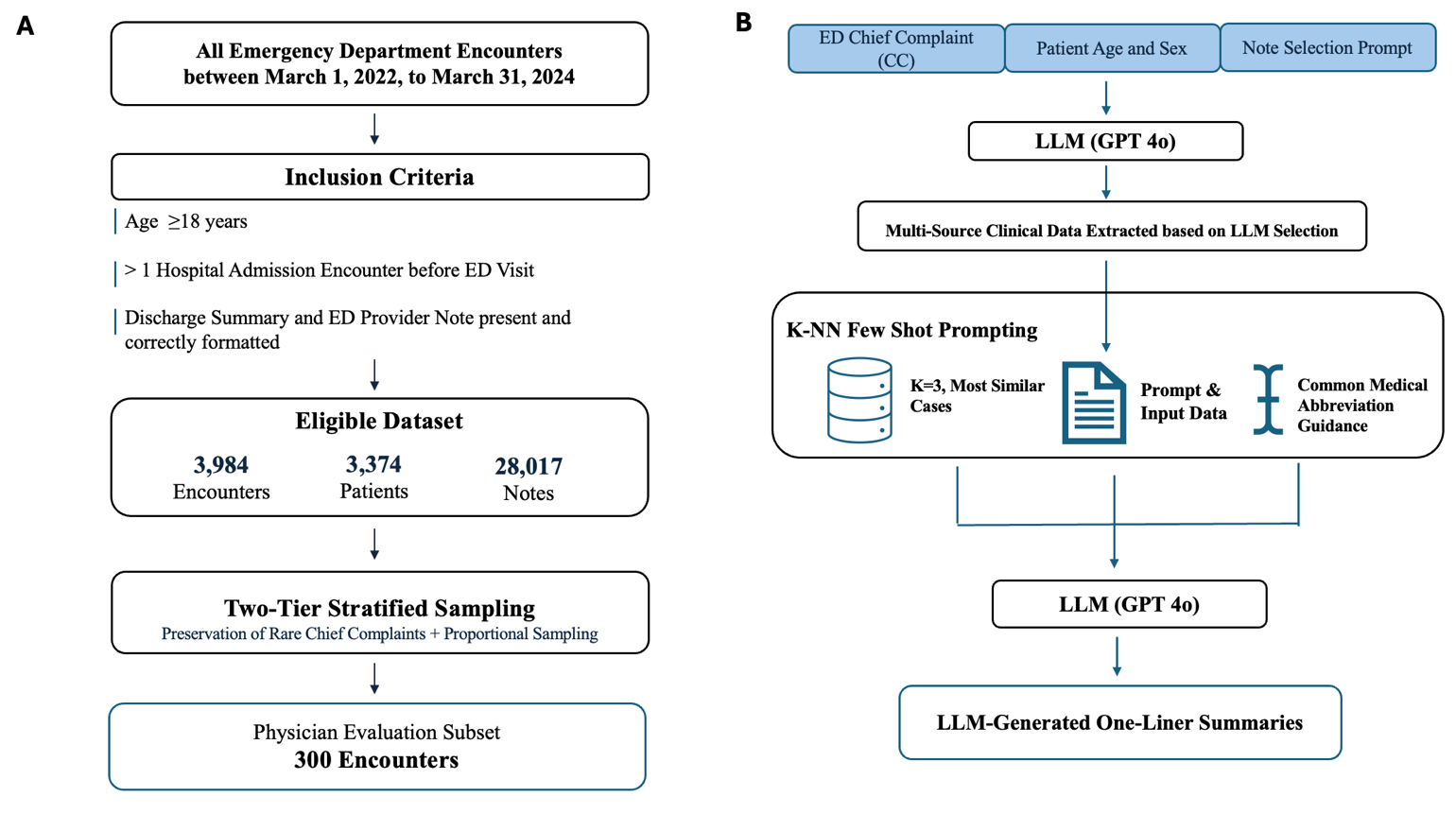
